# Respiratory Viral Contribution to Acute Myocardial Infarction: A Time Series and Spatiotemporal Analysis in Victoria, Australia 2010–2022

**DOI:** 10.64898/2026.01.30.26345252

**Authors:** Tu Q. Nguyen, Eric Zhao, Aaron L. Weinman, Benjamin D. Atkins, Tim Spelman, Suzanne Mavoa, Hazel J. Clothier, Christopher M. Reid, Jim P. Buttery, the SnotWatch Collaboration Group

**Affiliations:** Department of Paediatrics, University of Melbourne, Parkville, VIC, Australia; Centre for Health Analytics and Epidemiology-Informatics Research Group, Murdoch Children’s Research Institute, Parkville, VIC, Australia; Burnet Institute, Melbourne, VIC, Australia; Adelaide Medical School, University of Adelaide, Adelaide, SA, Australia; Centre for Health Services Research, Peter MacCallum Cancer Centre, Melbourne, VIC, Australia; Prevention Innovation, Murdoch Children’s Research Institute, Parkville, VIC, Australia; School of Population and Global Health, University of Melbourne, Parkville, VIC, Australia; School of Population Health, Curtin University, Bentley, WA, Australia; Centre for Cardiovascular Research and Education in Therapeutics, Monash University, Melbourne, VIC, Australia; Department of Infectious Diseases, Royal Children’s Hospital, Parkville, VIC, Australia

**Keywords:** myocardial infarction, respiratory viral triggers, influenza, spatiotemporal, vaccination

## Abstract

**BACKGROUND:** Respiratory viral infections can trigger acute myocardial infarction (AMI). However, the proportion of AMI events attributable to viral respiratory pathogens is unclear.

**METHODS:** This ecological study used time-series and spatiotemporal analyses to examine population-level patterns in Victoria, Australia, from 2010 to 2022. Independent statewide admissions and laboratory datasets were obtained. Generalized additive modelling was used to analyze the temporal association between respiratory viral circulation captured by polymerase chain reaction (PCR) testing and weekly counts of AMI admissions. A Bayesian hierarchical model was used to explore spatiotemporal variation in AMI associated with respiratory viruses.

**RESULTS:** Our study included 164 283 AMI hospital admissions and 6 180 896 PCR-tested samples. An increase in any respiratory virus detection rate was significantly associated with an increased incidence of AMI (incidence rate ratio [IRR] 1.0041; 95% confidence interval 1.0015–1.0067), after adjusting for seasonality, cold temperature, and fine particulate matter air pollution. An estimated 8.7% of total AMI events may be attributable to respiratory viral triggers, constituting an average annual incidence of 16.2 per 100,000 population. Significant associations were found with specific respiratory viruses; the fractions of AMI attributable to enterovirus, influenza, and respiratory syncytial virus were 5.2%, 1.5%, and 0.9%, respectively, with figures increased during peak viral seasons. Spatiotemporal analysis revealed that the association was more pronounced in outer-metropolitan areas.

**CONCLUSIONS:** Respiratory viral triggers contribute to the incidence of AMI. Population-level infection prevention strategies, such as vaccination, may reduce the impact of respiratory viral outbreaks during peak seasons.

**CLINICAL PERSPECTIVE:** *What Is New?:*

- Using time-series analysis and modern spatiotemporal techniques, we analyzed data from Victoria, Australia, to model population-level associations between AMI and respiratory viral activity and found that a recent laboratory-confirmed respiratory viral infection is associated with a higher incidence of AMI.
- An estimated 8.7% of total AMI events may be attributable to respiratory viral triggers, constituting an average annual incidence of 16.2 per 100,000 population.

*What Are the Clinical Implications?:*

- Some respiratory viral infections temporarily increase the risk of acute MI.
- With the existing vaccines available against influenza and respiratory syncytial virus (RSV), public health policy actions for influenza and RSV vaccination, particularly in high-growth urban areas, may help reduce the acute cardiovascular burden and health system strain.

## INTRODUCTION

Ischemic heart disease (IHD) is the leading cause of death worldwide.^1^ According to Global Burden of Disease Study estimates, IHD consistently ranks as the top contributor to global disease burden, accounting for 193 million (95% uncertainty interval 176–209) disability-adjusted life years across all ages and sexes.^2^ The most common acute clinical manifestation is acute myocardial infarction (AMI), a life-threatening event usually occurring when there is a sudden blockage in coronary artery blood flow.^3^ While the major risk factors are well-established, AMI is often preceded by precipitating risk factors, such as physical exertion, stressful events, drug use, air pollution and infections.^4–6^

Previous research has frequently implicated respiratory viruses, particularly influenza, as important precipitants of AMI.^7–9^ The incidence of AMI shows strong temporal associations and seasonal patterns, similar to those observed for influenza.^10^ The biological mechanisms are thought to be related to a transient pro-inflammatory, pro-coagulatory state in the acute phase response during an infectious episode, which can predispose individuals to ischemia and thrombosis within atherosclerotic coronary arteries.^11,12^ In a time-series analysis, García-Lledó et al^7^ found that an influenza-like illness rate of 50 per 100,000 population resulted in an increased risk of AMI during the same week (risk ratio 1.16, 95% CI 1.09–1.23), with the effect disappearing after one week. Similar observational evidence suggests that SARS-CoV-2, enterovirus, and respiratory syncytial virus (RSV) may also act as triggers,^13^ particularly over a short-term, transient risk period following infection.^4^ Respiratory viral infections is now recognized as an important driver of the atherosclerotic events due to their widespread prevalence in the community.^8,14^

Given the substantial contribution of AMI to the overall cardiovascular burden, randomized controlled evidence that influenza vaccines may reduce AMI risk,^15^ and growing evidence of the role of several common respiratory viruses,^13^ understanding the extent to which these infections may contribute to AMI events is urgently needed. Knowledge of the proportion of AMI attributable to respiratory viral pathogens, real-world dynamics, and the potential for risk reduction may be used to inform whether preventive approaches such as vaccinations are appropriate.^16^ Existing studies do not address attributable fraction measures as there is insufficient representative exposure data with which to reliably analyze the relationship. Laboratory surveillance data typically significantly under-represent actual numbers of infections in the community.^17^ Environmental epidemiological studies have assessed the impact of other less “proximal” risk factors such as temperature and air pollution on global cardiovascular burden.^18,19^ Meanwhile, there is minimal data available regarding attributable proportion for infections – the studies that do exist used UK Biobank data.^9,20^ These studies showed that 0.4%–3.0% of admissions among older persons (≥65 years) may be attributable to respiratory infections, with increased burden up to 7.7% when viral circulation is high,^9^ which implies that the largest impact may be observed during peak seasons. As infections usually exhibit substantial spatial clustering,^21^ we postulated that local outbreaks might influence AMI patterns when observed in the same time and space. If so, this would provide theoretical support for the exposure-response relationship used to generate attributable risk estimates.^22^

In this study, we aimed to investigate the respiratory viral contribution to AMI at a population level over a 13-year study period in Victoria, Australia. Our primary objective was to examine the incidence of AMI associated with respiratory viral triggers in a time-series analysis and to estimate the attributable fraction from this model. We considered temperature and ambient air pollution, particularly inhalation of fine particulate matter,^23^ as population-level confounders. A secondary objective was to explore potential space-time interactions between respiratory viral circulation and AMI incidence in a spatiotemporal analysis.

## METHODS

### Study Design and Setting

Victoria, an Australian state with a population of 6.5 million as of 2021, covers an area of 227,600 km^2^ (comparable in size to the UK), and represents about a quarter of the country’s population.^24^ The winter respiratory virus season typically spans from June to September. This study used ecological time series data from Victoria between 2010 and 2022. We analyzed laboratory-confirmatory testing data for respiratory viral infections, together with statewide hospitalization for AMI. Our analysis covered the period from June 1, 2010, to December 31, 2022. Data were obtained from January 1, 2010, but unavailability of electronic pathology records meant that laboratory data were incomplete prior to May 25, 2010; therefore, the study period was shortened to ensure temporal alignment between the main exposure and outcome datasets. A weekly temporal unit was the primary timescale of interest, based on previously documented risk windows for AMI and laboratory-confirmed respiratory viral infections.^9,25^

### Data Sources and Measures

Primary outcome AMI events were obtained from the Victorian Admitted Episode Dataset, a statewide administrative dataset that collects data on all hospital admitted episodes in public and private hospitals in Victoria. Deidentified unit record-level data in the form of International Statistical Classification of Diseases, Tenth Revision, Australian Modification (ICD-10-AM) codes for all admitted episodes within the study period were obtained from the Victorian Department of Health. Incident cases were defined by ICD-10-AM codes for AMI (I21.0–I21.9) as the principal (first) diagnoses, including ST-segment-elevation myocardial infarction (STEMI) and non-STEMI cases. Prior validation in ICD-10-AM has demonstrated sensitivity and specificity of the principal diagnosis field for AMI subtypes suitable for temporal trend analysis, rather than being affected by changes in coding over time.^26^ Patients with subsequent myocardial infarction or complications following AMI were excluded (**Methods S1**). The data extract included hospital admission date taken as the date of the event and patient location geocoded from usual residential address.

Respiratory viral circulation levels were determined from polymerase chain reaction (PCR) assays performed on respiratory specimens captured in “SnotWatch”; the methodology has been detailed previously.^27,28^ In brief, the SnotWatch collaborative data platform captures the results of specimens tested for routine clinical care or outbreak investigations performed by major diagnostic laboratories across Victoria (n = 7). These specimen-level data represent infectious patient episodes rather than individual persons. Data variables included specimen collection date, postcode of usual patient residence, and pathogen-specific results (e.g., detected, not detected). We analyzed PCR results for respiratory viral targets: adenovirus, bocavirus, seasonal coronaviruses, enterovirus (including rhinovirus species), human metapneumovirus (hMPV), influenza, parainfluenza virus 1-4, RSV, and SARS-CoV-2 (COVID-19), using data available as of September 2025. To account for differences between individual laboratories with different assays and viral targets, laboratory-specific data were cleaned and standardized separately before being combined and aggregated (**Methods S2**). For this analysis, we calculated the test positivity rate as a proxy measure for circulating respiratory virus levels, expressed as the proportion of positive detections for each pathogen: *v_t_ = p_v_ / total_v_*, where *v_t_* denotes the exposure level within weekly time interval *t,* calculated by dividing the number of positive specimens (*p_v_)* by the total number of specimens tested for virus *v (total_v_)*. The specimen collection date was used to aggregate weekly test positivity. We used test positivity instead of the absolute number of positive detections to compensate for changes in laboratory testing practices over time.^29^

In terms of covariates, weather data were obtained from the Australian Bureau of Meteorology for all stations in Victoria (n = 109), including daily minimum temperature (°C)^30^ and relative humidity (% of the amount of moisture the air can contain). Daily data were aggregated into weekly average (mean) minimum temperature. Ambient air pollution estimates were obtained from the National Air Pollution Monitoring Database,^31^ specifically daily fine particulate matter ≤2.5μm (PM_2.5_) data from all monitoring stations in Victoria, from which weekly mean concentrations were estimated. Population denominator data were downloaded from publicly available Australian Bureau of Statistics (ABS) estimated resident population (ERP) as per 2021 Census.^32^ The ERP is the official estimate of the population published by the ABS and is released annually (June 30) at the sub-state level. Given the short (weekly) time unit of interest, we assumed a constant population throughout the year; that is, it was relatively stable from week to week. We used ERP to calculate the incidence of AMI per 100,000 person-weeks.

For the secondary objective, we expanded the time series to a spatiotemporal dataset comprising weekly data split by Australian Statistical Geographical Standard (ASGS) Statistical Areas Level 2 (SA2) spatial units, the finest spatial resolution publicly available for resident population estimates.^33^ SA2 is a medium-sized area that represents an economic and social community with (population of approximately 10,000 people). According to 2021 classifications, Victoria had 524 SA2 regions; our analysis covered 520 unique SA2s, after excluding zero population regions. Location in the hospital admissions data was by SA2 region. To align spatial units, we mapped residential postcodes from respiratory PCR data to SA2 regions using publicly available concordance tables.^33,34^

### Data Analysis

We summarized the mean (standard deviation [SD]) and median (interquartile range [IQR]) values of continuous variables. Frequencies and percentages were used to summarize the categorical variables.

#### Time-Series Analysis

We first analyzed the temporal association between AMI counts and short-term changes in respiratory viral test positivity in a time-series analysis. A generalized additive model (GAM) with a log link was applied following a negative binomial distribution. This was used instead of a Poisson distribution due to overdispersion in the count data.^35^ To account for seasonality and long-term trends, we used spline functions to allow smooth modelling over time. A thin plate regression spline of the weekly index for time (1:657) was used; this ensured that seasonal patterns were pathogen-specific, as different respiratory viral pathogens have clearer cyclical patterns and winter peaks than others.^36^ Minimum temperature and PM_2.5,_ were considered as covariates. To account for changes in laboratory testing behavior associated with the COVID-19 pandemic, we explored the inclusion of a binary variable for pre- and post-2020 as an interaction term with viral exposure. Given the long study period, we included a population offset to control for the population size changing over time. The model may be expressed as: *AMI ∼ Virus_t-lag_ + spline + O_t_ + MinTemp + PM_2.5_ ± COVID-19*; where *AMI* denotes the count of AMI admissions, *Virus_t-lag_* is virus positivity (%) at week *t-lag*, *spline* is the thin plate regression spline; and *O_t_* is the annual ERP count in log scale as an offset. Model coefficients were exponentiated to derive incidence rate ratios (IRR) with 95% confidence intervals calculated using the Wald method. In the presence of population offset *O_t_*, this was interpreted as the multiplicative change with respect to the mean AMI incidence rate.

Prior studies have shown that exposure to respiratory viral infection increases the short-term risk of AMI for a period lasting several days to weeks following a laboratory confirmatory test.^13^ To account for potential temporal lag-response associations, we allowed for lag-lead effects of within 3 weeks of exposure, with outcome shifted accordingly (*lag* = ±3 weeks) – positive lags allow a delay between the exposure and its subsequent effect on outcome whereas negative lags (lead times) is when the exposure appears to increase/decrease after the outcome effects have already been observed. Similar to previous studies,^7,9^ potential lag times were examined to account for the time between infection and AMI onset, while lead times were used as laboratory surveillance generally reflects respiratory viral circulation in the preceding weeks.^37^ Sensitivity analyses examined: seasonal adjustment methods (time-stratified vs. spline functions); addition of covariates; impact of pre- and post-COVID period; alternative temporal units (daily, monthly, annual); and wider temporal lagged periods. Models were calibrated using the Akaike information criterion/Bayesian information criterion.^38^

Subsequently, a distributed lag non-linear model (DLNM) was used to compute attributable risk from the ‘net’ effect over all lags.^39^ Lag-lead times were modelled by replacing the single lagged variable *Virus_t-lag_* from the above equation with a cross-basis function of tensor product smooth over lag *cb (Virus_t_, lag)*. Confounders were not lagged as PM_2.5_ and low temperatures have relatively short-lived effects and it is unlikely to have the same lagged effects of up to 3 weeks as in respiratory viral circulation.^40,41^ Model coefficients from the fitted DLNM were used to compute predicted associations of the total number of AMI cases attributable with effects distributed over the full lag period.^42^ The model-based population attributable fraction (PAF) was estimated by predicting the AMI incidence rate under observed viral circulation *V_t_* compared to a counterfactual scenario of zero circulating virus for each observed week *t* in the study period. The PAF was calculated as: *PAF = Σ_t_ AN(V_t_) / Σ_t_ y_t_;* representing the fraction of all AMI attributable to non-zero viral circulation derived from the weekly mean computed totals. PAF was then estimated by peak and low viral seasons, defined as the 90^th^ and 10^th^ percentiles of weekly test positivity levels, respectively. We calculated the annual number of cases and annual incidence per 100,000 population by dividing the mean annual attributable cases by the mean ERP. The 95% confidence intervals were derived from the 2.5^th^ and 97.5^th^ percentiles of the simulated distribution constructed from 1000 Monte Carlo simulations. Analyses were performed using R Statistical Software (version 4.4.3, packages “mgcv”, “splines”).^43^

#### Spatiotemporal Analysis

As a descriptive step, we calculated Moran’s *I* statistic to identify global and localized spatial correlation at the SA2-level for AMI cases across Victoria.^44^ To explore a potential spatiotemporal association between AMI and viral circulation, a Bayesian hierarchical approach was used, modelled over a space-time index that identifies both time point and the associated spatial location.^45^ We initially sought to add a tensor product smooth for space-time interaction to the time-series model; however, the GAM was challenging to model robustly due to low cell counts across SA2-weeks, resulting in unstable estimates. Hence, a Bayesian method was preferred due to being particularly effective and flexible enough to apply to complex data structures while providing reliable quantification of uncertainty.^46,47^

The spatiotemporal model was specified using an Integrated Nested Laplace Approximation (INLA) for marginal (posterior) inference, following established methods.^37,46,47^ Like the time-series, weekly AMI counts were assumed to follow a negative binomial distribution, and ERP was included as an offset. The model included a fixed effect for viral activity and random effects to account for spatial, temporal, and space-time processes at the SA2-week level. Spatial neighborhood adjacency structure used ASGS SA2 boundaries (**Methods S3**). Spatial variability was modelled using a Besag–York–Mollié (BYM) specification, comprising two components: (1) a structured residual with a conditional autoregressive model constructed as a function of first-order neighborhood to enable AMI rates to be influenced by viral activity from neighboring SA2 regions, and (2) an unstructured residual to allow independent spatial heterogeneity not explained by covariates. Temporal trends were modelled using a first-order autoregressive process over weeks (1:657). Additional variability between SA2-weeks was captured using an unstructured space–time interaction and a first-order random walk to allow smooth temporal evolution. An SA2-specific random effect for viral activity was included, which enabled estimation of differential effects in respiratory viral triggers beyond the average association. Default priors were used for the fixed effects and hyperparameters. Spatial, temporal, and space-time random effects were specified with penalized complexity priors on precision parameters. Environmental covariates were not included due to lack of (readily available) spatially resolved data at a weekly resolution. We exponentiated the posterior means of the coefficients for the fixed effect of viral exposure to derive IRRs with 95% credible intervals. Model were calibrated using the Watanabe–Akaike information criterion and deviance information criterion. Sensitivity analyses were conducted on alternative model structures to assess robustness to the specifications. Further model details are in **Methods S3**.^48^ We used R packages “spdep”, “INLA”, “sf”.^46,47^

### Ethics

This study was conducted with approval from Monash Health Human Research Ethics Committee (Reference: RES-19-0000333L-53611) and the University of Melbourne Office of Research Ethics and Integrity (Reference: 2024-28658-48562-1). A waiver of the requirement for individual consent was obtained for this study. Reporting adheres to the Strengthening the Reporting of Observational Studies in Epidemiology (STROBE) guidelines (**Supplementary File 2**).^49^

## RESULTS

### Descriptive Statistics

The key descriptive statistics of AMI cases, viral circulation and environmental factors are summarized in Table 1. A total of 164 283 AMI cases in Victoria were admitted between June 1, 2010, and December 31, 2022 (66.4% male; 63.1% aged ≥65 years; 25.1% STEMI). On a weekly basis, AMI cases had a mean (SD) of 250.0 (27.5) per week. There was evidence of winter peaks in hospital admissions for AMI, whereas long-term trends were relatively stable over the study period (**Figure 1**). Over the same period, 6 180 896 respiratory PCR-tested specimens were recorded, of which 664 626 (10.7%) returned positive results for any virus. Although the number of PCR samples was highly variable from week to week, including notable shifts during the COVID-19 pandemic, strong seasonality was evident from respiratory virus test positivity, particularly for influenza, RSV, and enteroviruses (including rhinovirus species).

**Figure 1.**
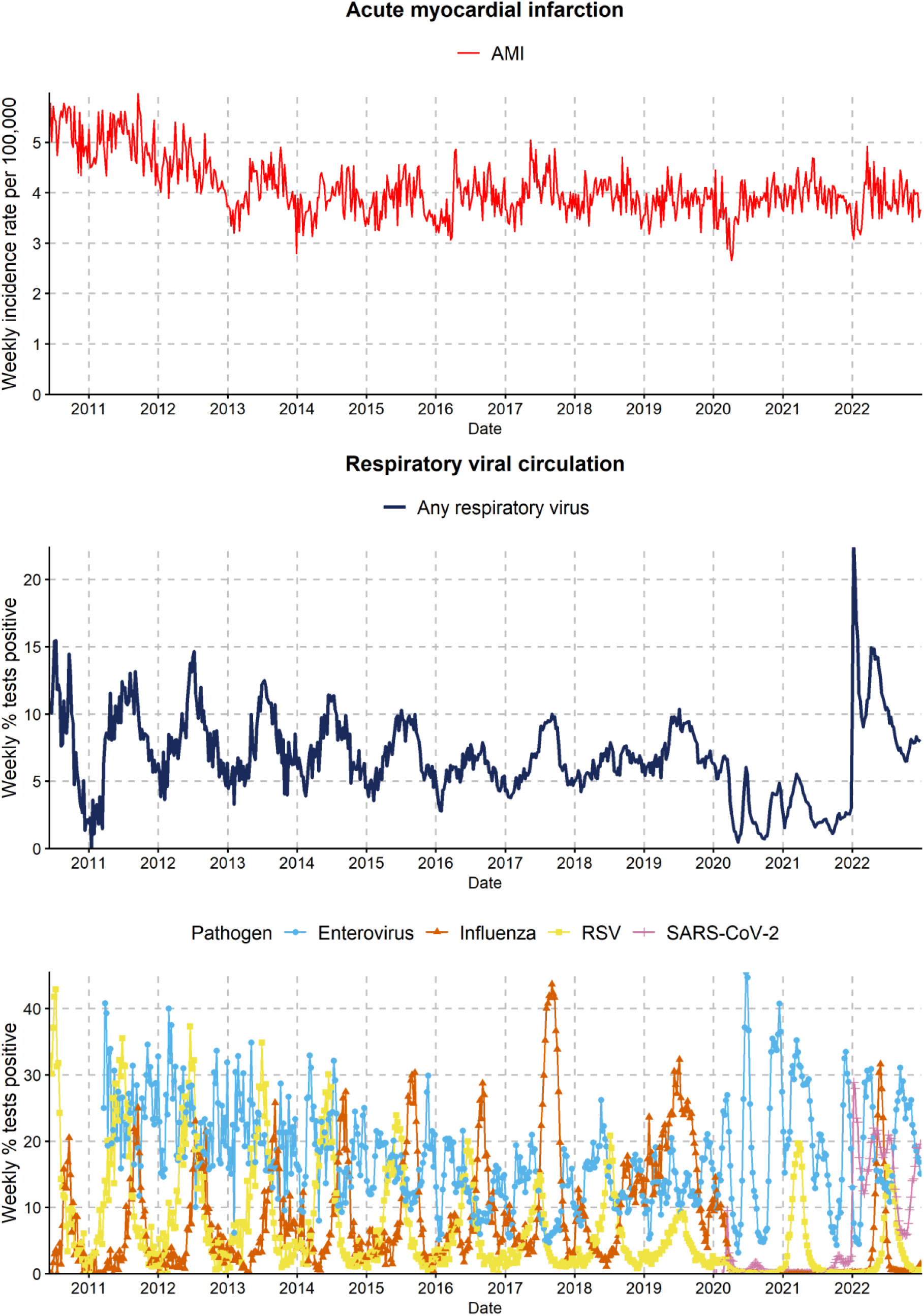
Weekly Time Series of AMI and Respiratory Viral Circulation Levels in Victoria, 2010 to 2022. Abbreviations: AMI, acute myocardial infarction; RSV, respiratory syncytial virus.

**Table 1.** Descriptive Statistics of AMI, Respiratory Viral Circulation, and Environmental Factors in Victoria, 2010 to 2022.

|  | Total, N | Weekly mean (SD) | Weekly median (IQR)<br>[range] |
| --- | --- | --- | --- |
| **AMI** |  |  |  |
| No. of cases | 164 283 | 250 (28) | 251 (35) [165-331] |
| Incidence rate per 100,000* | N/A | 4.1 (0.6) | 4.0 (0.6) [2.7-6.0] |
| **Respiratory viral circulation** |  |  |  |
| Total specimens | 6 180 896 | 9397 (21 668) | 605 (3198) [26-128 617] |
| Test positivity (any virus), % | 10.7 | 6.8 (3.1) | 6.6 (3.6) [0-22.4] |
| **Environmental factors** |  |  |  |
| Low temperature, °C | N/A | 8.8 (3.8) | 8.3 (6.0) [2.3-18.6] |
| PM<sub>2.5</sub>, µg/m<sup>3</sup> | N/A | 6.5 (3.9) | 5.9 (2.6) [1.7-75.1] |

Abbreviations: AMI, acute myocardial infarction; IQR, interquartile range; N/A, not applicable; PM<sub>2.5</sub>, particulate matter with diameter of 2.5 mm or less; SD, standard deviation.
\* Crude rate per 100,000 person-weeks.

### Temporal Association Between AMI and Respiratory Viruses

In the time-series analysis, the population incidence of AMI was significantly associated with an increase in test positivity for any respiratory virus (IRR 1.0041; 95% CI 1.0015–1.0067 at - 3 weeks lag). **Table 2** shows the temporal association between AMI and respiratory viral activity from single-lag models, after controlling for low temperature and PM_2.5_ concentration. In terms of specific pathogens, influenza, enterovirus, and RSV had significant effects over several single-lag periods, with the greatest effect sizes observed at - 3 weeks, -2 weeks, and 3 weeks lag, respectively. We found hMPV was only significant when lagged at -1 week (IRR 1.0031; 95% CI 1.0007–1.0054). There was no evidence of a temporal association for SARS-CoV-2, adenovirus, parainfluenza virus 1-4, bocavirus, or seasonal coronaviruses. A clear time-varying pattern was observed, with reductions in risk observed at longer lags for any respiratory virus, particularly for enterovirus (**Figure S1**). Wider single lagged effects in sensitivity analysis revealed an inverted U-shaped dose-response relationship (**Figure S2)**, suggesting a broadly linear association for shorter lag periods from - 8 to +4 weeks, with the effects diminishing over time.

**Table 2.** Temporal Association Between AMI and Respiratory Viruses.

| Pathogen | Lag* | IRR (95% CI) | P value<sup>†</sup> |
| --- | --- | --- | --- |
| Any respiratory virus | -3 weeks | 1.0041 (1.0015–1.0067) | 0.002<sup>‡</sup> |
| Influenza | -3 weeks | 1.0011 (1.0004–1.0018) | 0.003<sup>‡</sup> |
| SARS-CoV-2 | -3 weeks | 0.9986 (0.9954–1.0018) | 0.391 |
| Enterovirus | -2 weeks | 1.0022 (1.0012–1.0032) | <0.001<sup>‡</sup> |
| RSV | 3 weeks | 1.0012 (1.0001–1.0023) | 0.027<sup>‡</sup> |
| Adenovirus | 0 weeks | 1.0018 (0.9988–1.0048) | 0.240 |
| Parainfluenza 1-4 | 0 weeks | 1.0016 (0.9985–1.0047) | 0.305 |
| hMPV | -1 week | 1.0031 (1.0007–1.0054) | 0.011<sup>‡</sup> |
| Bocavirus | -3 weeks | 1.0069 (0.9973–1.0167) | 0.163 |
| Seasonal coronavirus | -2 weeks | 0.9969 (0.9893–1.0046) | 0.427 |

Abbreviations: AMI, acute myocardial infarction; hMPV, human metapneumovirus; IRR, incidence rate ratio; RSV, respiratory syncytial virus.
\* Lag observed with the greatest effect size (IRR > 1) within 3 weeks.
† P value for lag-specific estimate adjusted for temporal trend, minimum temperature (°C), PM2.5 concentration and COVID-19 period (if applicable).
‡ Indicates significant differences (P < 0.05).

Sensitivity analysis demonstrated that our results were robust to alternative regression modelling, method of seasonal adjustment, alternative temporal units (on a daily and monthly basis), and adjustment for the COVID-19 pandemic (**Table S1**). While the best fitting models controlled for both low temperature and PM_2.5_ concentration, inclusion of low temperature had a more noticeable impact than PM_2.5_ on model estimates, suggesting temperature was a more important confounder to control for.

### Annual AMI Events Attributable to Respiratory Viral Triggers

**Table 3** shows the proportion of AMI events attributable to respiratory viral triggers based on the distributed lag time-series model. An estimated 8.69% (95% CI 4.59–12.64) of AMI events were attributable to any respiratory virus. Annually, the fraction attributable to any respiratory virus was equivalent to an annual average of 16.23 (95% CI 8.58–23.61) per 100,000 population. The fractions of AMI attributable to each virus were 5.16%, 1.47%, and 0.94% for enterovirus, influenza, and RSV, respectively. Once the cumulative association with hMPV was estimated over all lags, the association with AMI incidence was no longer significant. During peak viral seasons, the PAF for enterovirus rose to 9.21%, the highest pathogen-specific estimate, followed by RSV at 3.37% and influenza at 2.85%. Due to the composite effect of different pathogens, an increase during the peak viral season was not observed with any respiratory virus. On average, the annual AMI incidence attributable to each pathogen was 8.95 (95% CI 5.06–11.18) for enterovirus, 2.74 (95% CI 0.50–5.05) for influenza and 1.75 (95% CI 0.25–3.11) for RSV per 100,000.

**Table 3.** AMI Events Attributed to Respiratory Viral Triggers, Fraction and Average Annual AMI events*.

| Pathogen<sup>‡</sup> | Fraction attributable, % (95% CI) |  |  | Annual AMI events, mean (95% CI) |  |
| --- | --- | --- | --- | --- | --- |
|  | Overall | Low season<sup>†</sup> | Peak season<sup>†</sup> | No. of cases | Incidence per 100,000 |
| Any respiratory virus | 8.69 (4.59 to 12.64) | 2.54 (0.72 to 4.33) | 8.46 (4.53 to 12.60) | 997 (527 to 1451) | 16.23 (8.58 to 23.61) |
| Influenza | 1.47 (0.27 to 2.70) | 0.00 (0.00 to 0.00) | 2.85 (0.011 to 5.54) | 168 (31 to 310) | 2.74 (0.50 to 5.05) |
| Enterovirus | 5.16 (2.91 to 7.43) | 1.93 (1.08 to 2.80) | 9.21 (5.23 to 13.16) | 550 (311 to 792) | 8.95 (5.06 to 12.88) |
| RSV | 0.94 (0.13 to 0.67) | 0.01 (0.00 to 0.02) | 3.37 (0.49 to 5.98) | 107 (15 to 191) | 1.75 (0.25 to 3.11) |
| hMPV | 0.56 (-0.44 to 1.57) | 0.00 (0.00 to 0.00) | 1.48 (-1.19 to 4.15) | 64 (-51 to 180) | 1.04 (-0.83 to 2.93) |

Abbreviations: AMI, acute myocardial infarction; CI, confidence interval; hMPV, human metapneumovirus; PAF, population attributable fraction; RSV, respiratory syncytial virus.
\* Within confidence intervals, 'to' has been used because of negative values in the span.
<sup>†</sup> Low and peak defined as the 10<sup>th</sup> and 90<sup>th</sup> percentiles of weekly levels of circulating virus, respectively.
<sup>‡</sup> Specific respiratory viral pathogens were analyzed separately. Therefore, results were not additive and attributable fractions cannot be summed together.

### Spatiotemporal Differences Between AMI and Respiratory Viruses

**Figure 2** depicts the geographical distribution of respiratory viral test positivity and average weekly AMI incidence per 100,000 population across SA2 regions in Victoria. There was clear spatial heterogeneity in the respiratory virus levels, whereas AMI had no overall spatial correlation (Global Moran’s *I* statistic 0.0073; p = 0.58). There was, however, evidence of localized spatial clustering in AMI cases and those of nearby SA2 regions with Local Moran’s *I* statistic (**Figure S3**).

**Figure 2.**
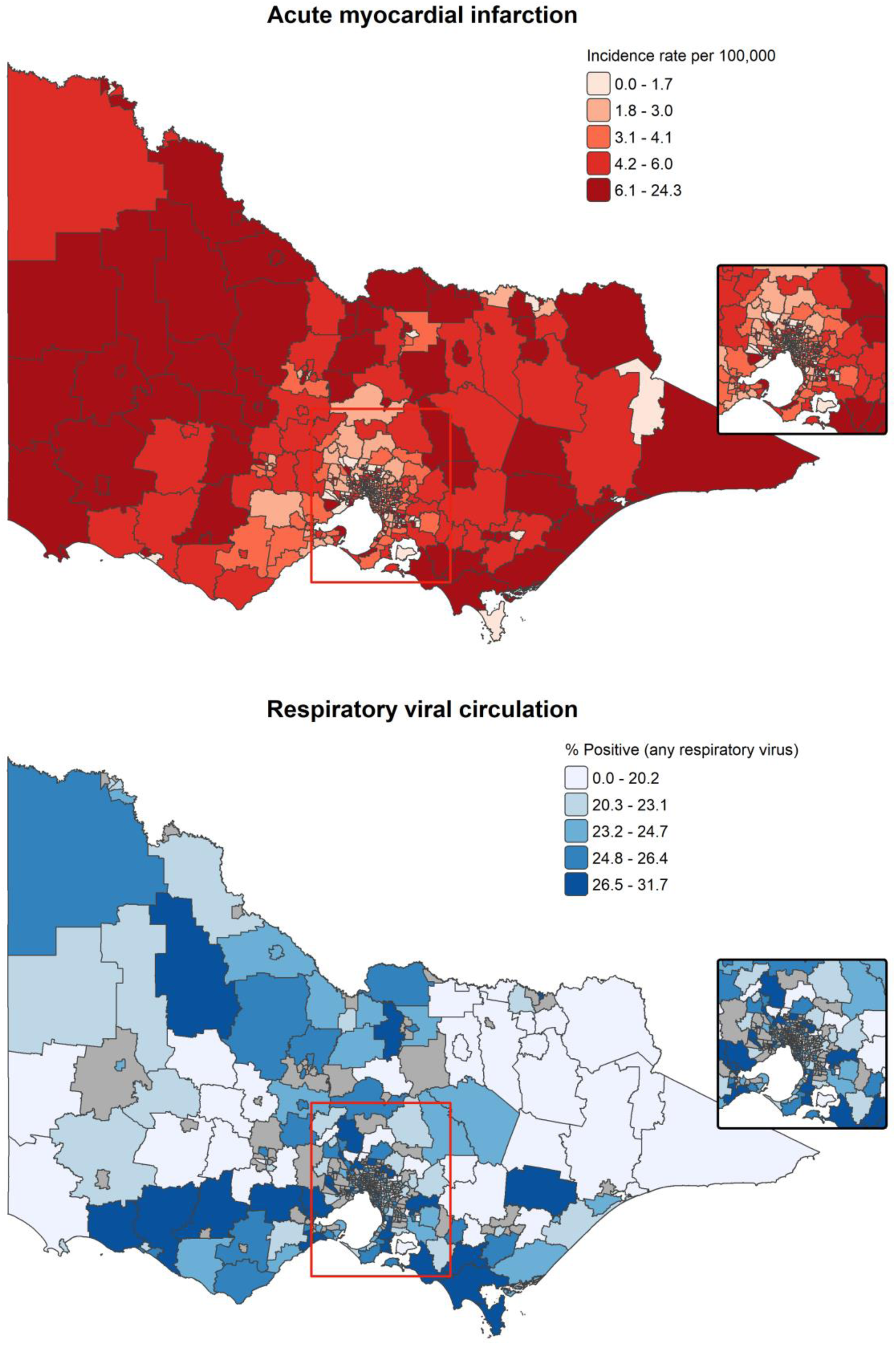
Geographic Distribution of Weekly Mean AMI Incidence Rate per 100,000 and Respiratory Viral Circulation Across Victoria and Metropolitan Melbourne. SA2 region-specific means over the entire study period. The inset map provides a detailed view of the Metropolitan Melbourne area. Abbreviations: AMI, acute myocardial infarction, SA2, Statistical Area Level 2.

In the spatiotemporal analysis, the incidence of AMI was significantly associated with increased respiratory viral levels at the SA2-week level (IRR 1.0015; 95% credible interval 1.0009–1.0021), after accounting for viral exposure in neighboring SA2s and across regions, temporal trend, and space-time interactions. While the time-series and spatiotemporal analyses were analyzed using different models and comparing them is not valid, the results were generally consistent. Comparable findings were observed for specific viruses, except RSV (**Table S2)**. Higher estimates of the differential viral space-time effects were predominantly from urban areas on the outskirts or just outside the capital city of Melbourne (**Figure S4**).

## DISCUSSION

Although respiratory viral infections have been increasingly recognized as important triggering factors for acute cardiovascular events,^8,9,20^ knowledge of their real-world contribution is scarce. Global estimates suggest that the burden of IHD will continuously increase until 2050,^50^ consistent with Australian modelling projections for AMI.^51^ Our aim was to understand the extent to which respiratory viruses contribute to AMI using large-scale population data over a 13-year study period. We estimated that approximately 8.7% (95% CI 4.6–12.6) of total AMI events may be attributed to respiratory viral infections, with the proportion rising during peak viral seasons, consistent with data from UK Biobank studies.^9,20^ As previous studies have not explored space-time interaction, we hypothesized an observable spatiotemporal association. Spatiotemporal analysis yielded results consistent with the traditional time-series model and provided further support for the population-level association.

Our results show that, after controlling for the effects of temporal trends, climate, and fine particulate matter air pollution, higher circulating levels of any respiratory viruses were associated with a small but incremental increase (of 0.41%) in the incidence of AMI. The time-series analysis aligns with prior studies reporting comparable effect sizes for increased risk within 2–3 weeks of infection.^7,9,20^ As in previous studies, our results reinforce the importance of timing.^13^ Analysis of single lagged effects suggested a time-varying pattern, with the greatest effect in the immediate short-term period that seems to diminish with recovery from infection over time, which supports the theory that acute infectious episodes may act as a catalyst for the progression of vulnerable plaques in the short-term.^52^ However, our results differ from those of self-controlled case series studies that identified the strongest positive associations in the first few days immediately following a positive test.^8,14^ We found longer temporal lag effects than those observed in other studies (typically ±2 weeks), suggesting delayed clinical diagnosis for infections and hence extended time for referral to laboratory surveillance.^9^ A possible reason for this might be that delays during outbreaks, due to prioritization of treating patients rather than reporting cases,^37^ are more pronounced in population-level data compared to individual-level data. Furthermore, an interesting finding was the sensitivity analysis on wider temporal lags which showed an apparently protective effect at longer lags, potentially due to depletion of the susceptible pool.^40^ This confirmed the need for a distributed lag model to assess the cumulative effect over the full lag period of interest.

We specifically identified the contribution of RSV, influenza, and enterovirus to be between 0.94%–5.16%. The biological mechanisms of respiratory viral triggers for AMI have been suggested to be related to inflammatory or immune responses. These effects are not thought to be pathogen-specific, although respiratory viruses entering the systemic circulation may invoke different pro-inflammatory responses.^53^ Contrary to expectations, our results did not indicate an association between SARS-CoV-2 and AMI admissions. This finding may reflect either a true lack of association or insufficient time elapsed since 2020 to discern significant trends, although this is difficult to discern given the inconsistent evidence in existing literature.^13^ Whereas previous literature has consistently reported that the highest burden for AMI admissions was attributable to influenza,^13,14^ enterovirus had the highest estimates in our analysis. High detection rates in our exposure data could be a factor. Historically, locally used laboratory PCR panels have been unable to differentiate from within the *Picornaviridae* family, particularly enteroviruses. Rhinovirus is classified within the enterovirus genus but is considered distinct from other enteroviruses due to different clinical manifestation and seasonal pattern,^54^ therefore grouping all enteroviruses together may have produced higher estimates than rhinovirus alone. As it was not possible to differentiate between specific enteroviruses, further investigation with more specific tests is needed.

Prior analyses have primarily focused on the time-varying effects of respiratory viral triggers,^7,9,55^ with limited spatial analyses albeit space-time interactions. We found that the incidence of AMI was spatiotemporally associated with increased viral levels (IRR 1.0015; 95% credible interval 1.0009–1.0021), after accounting for viral exposure in neighboring regions and space-time interactions at the SA2-week level. Interestingly, there were differential viral space-time effects in some regions above the average association, suggesting spatial heterogeneity in the impact of respiratory virus circulation on AMI incidence beyond the baseline spatial and temporal trends. There were notably higher estimates in urban fringe areas and on the outskirts of Melbourne, which is consistent with “growth corridor” areas experiencing high population growth.^32^ The observed variation may reflect patterns of cardiovascular risk factors, such as prevalence of uncontrolled elevated blood pressure, differential healthcare access, region-specific age structure and socioeconomic status.^56^ We did not explore socioeconomic status because socioeconomic indices for this type of analysis would likely exhibit collinearity at the SA2-level.^57^ There was also potential for residual confounding, since we did not control for environmental covariates. Furthermore, vaccination rates against seasonal influenza and RSV may differ between geographic areas,^58^ but assessing this variation is challenging without reliable vaccine coverage data for the population. Although this should be interpreted as spatial heterogeneity rather than causal geographic effects, our findings support the theory that localized respiratory viral outbreaks may explain some variation in AMI, especially in similarly developed countries with temperate climates.

### Strengths and Limitations

The strengths of this study include the large, representative sample size over a 13-year study period and use of population-based data sources. Our exposure dataset is likely to be broader and more comprehensive than laboratory surveillance data based on notifiable disease cases alone.^36^ In the time-series model, we employed noise-reducing strategies to adjust for long-term fluctuations and population-level confounders including temperature and ambient air pollution, whereas prior studies had only controlled for climactic factors.^7,9,55^ We thoroughly assessed the effects of temporal lags, including cumulative risk over lag periods to compute attributable risk estimates, and supplemented the traditional time-series analysis with modern spatiotemporal techniques to support our assumptions.

This study had several limitations. First, this was an ecological study with reference to population-level information, and the findings cannot be used to make individual-level causal inferences. For example, using hospital admitted episodes to analyze AMI incidence cannot account for preexisting cardiovascular risk profile, potentially overestimating risk attributable to respiratory viruses. The dataset and covariates are inherently ecological, and the same inferences cannot be made as with individual data points. Second, we focused on respiratory viruses, which have been most frequently cited in previous studies and are the most common identified infectious agents in laboratory PCR testing. It is reasonable to assume that our results underestimate the proportion of AMI attributable to all infections, as bacterial pathogens, urinary tract infections, and gastrointestinal infections may also contribute to AMI incidence.^20^ Third, the spatiotemporal analysis was based on the SA2 region of usual residential address. This is likely to capture the viral circulation resulting from movements within a week in one’s residential area and neighboring regions, but not extending to more distant locations, such as workplaces. We also cannot exclude the ‘modifiable spatial unit problem’ whereby conclusions may differ if conducted at a finer scale.^59^ Further studies, ideally with finer spatial resolution e.g. local government areas, and spatially resolved covariate data are needed to meaningfully analyze and interpret the spatiotemporal association.

### Implications of Findings

Respiratory viral triggers were not analyzed with the full spectrum of cardiovascular risk factors; hence, the attributable fraction estimates should be interpreted accordingly. We specifically identified the contribution of enterovirus, influenza, and RSV. With the existing vaccines available against influenza and RSV, public health policy actions for influenza and RSV vaccination, particularly in high-growth urban areas, may help reduce the acute cardiovascular burden. On the other hand, absolute risk-reduction approaches to cardiovascular disease prevention have been shown to be more cost-effective overall, rather than addressing risk factors individually.^60^ Given our findings, future scenario-based and cost-effectiveness modelling should include infections analyzed in conjunction with the traditionally recognized risk factors to inform policy development around preventable cardiovascular outcomes. Considering the burden of AMI globally and in Australia,^61^ it is likely that even a modest decrease in infection-attributable risk would produce a net reduction in costs to the health system.

## CONCLUSION

Our results showed that respiratory viral infections contribute to AMI events. We specifically identified the contribution of enterovirus, influenza, and RSV. Our findings suggest that the incidence of respiratory virus-attributable AMI appears to vary by the timing and location of viral outbreaks. Public health policy actions for the prevention and control of infections, especially during peak periods, may help reduce the acute cardiovascular burden.

## Data Availability

The analysis R code is stored in an institutional Gitlab repository and is available from the corresponding author upon reasonable request. The NAPMD data is free available for research upon request to CARDAT. Climate data can be obtained directly from the Bureau of Meteorology under license. The VAED and SnotWatch data cannot be made publicly available and requires appropriate ethics and data custodian approvals.

http://www.bom.gov.au/climate/data/

https://cardat.github.io/napmdtools/using-napmd.html

## Nonstandard Abbreviations and Acronyms

ABS: Australian Bureau of Statistics
AMI: Acute myocardial infarction
ASGS: Australian Statistical Geographical Standard
DLNM: Distributed lag non-linear model
ERP: Estimated resident population
GAM: Generalized additive model
hMPV: Human metapneumovirus
ICD-10-AM: International Statistical Classification of Diseases, Tenth Revision, Australian Modification
IHD: Ischemic heart disease
IRR: Incidence rate ratio
INLA: Integrated Nested Laplace Approximation
PAF: Population attributable fraction
PCR: Polymerase chain reaction
PM_2.5_: Particulate matter with diameter of 2.5 mm or less
RSV: Respiratory syncytial virus
SA2: Statistical Areas Level 2
STEMI: ST-segment elevation MI

## *SnotWatch Collaboration Group

Monash Pathology (Tony Korman), Royal Children’s Hospital Pathology (Andrew Daley, Vanessa Clifford), Alfred Pathology (Adam Jenney), Royal Melbourne Hospital Pathology (Katherine Bond), Eastern Health Pathology (Roy Chean), Northern Pathology Victoria (Yvonne Hersusianto), Barwon Health (Eugene Athan), Victorian Department of Health (Jim Black).

## ACKNOWLEDGEMENTS

Thank you to all SnotWatch laboratories, investigators and their staff for provision of laboratory data used in this study; Li-yin Goh (Royal Children’s Hospital) for assistance with obtaining RCH laboratory data; the Victorian Department of Health for provision of VAED data; Daneeta Hennessy (Murdoch Children’s Research Institute) and Janet Strachan (Victorian Department of Health) for general epidemiological advice; and the University of Melbourne Statistical Consulting Centre, particularly Jeremy Silver, for advice on statistical methods and training. We are grateful to the clinical coding teams at the Royal Melbourne Hospital and Royal Children’s Hospital, especially Kyle Batty, Megan Christison and Allison Lusher, for advice on ICD-10 coding of cardiovascular conditions. This research was undertaken with the assistance of resources from the Clean Air Research Data and Analysis Tools (CARDAT) platform (https://cardat.github.io). CARDAT is supported by The Centre for Safe Air (https://safeair.org.au/), which is funded by the National Health and Medical Research Council (2015584) and the Australian Research Data Commons (ARDC) AirHealth Data Bridges project (https://doi.org/10.47486/PS022), and the WHO Collaborating Centre for Climate Change and Health Impact Assessment.

## SOURCES OF FUNDING

Tu Q. Nguyen was supported by an Australian Government Research Training Program PhD Scholarship and a Murdoch Children’s Research Institute Top-Up Scholarship during the preparation of this manuscript. Suzanne Mavoa was a GenV Fellow and supported by a FAIR Fellowship 2024 Award administered by veski for the Victorian Health and Medical Research Workforce Action Plan on behalf of the Victorian Government; funding for the Award has been provided by the Victorian Department of Jobs, Skills, Industry and Regions. Research at the Murdoch Children’s Research Institute was supported by the Victorian Government’s Operational Infrastructure Program. The SnotWatch project was supported by the Royal Children’s Hospital Foundation (Buttery Research Grant). Funding sources had no role in the development of the protocol, conduct of the systematic review, writing up, or decision to submit for publication. The funders had no role in study design, data collection, data analysis, data interpretation, writing up, or decision to submit for publication.

## DISCLOSURES

Jim P. Buttery reports participation on a data and safety monitoring board for the Coalition for Epidemic Preparedness and Innovation, academic institutions, and GSK, for which his employing institution (Murdoch Children’s Research Institute) is compensated for his time, as well as financial support for serving on the Australian Therapeutic Goods Administration Advisory Committee on Vaccines, outside the submitted work. Tim Spelman reports receiving compensation for serving on advisory boards and scientific committees from Biogen and Novartis, and consultancies from Abbott and Otsuka. All other authors report no potential conflicts.

## Notes

### Clinical Trial

Not applicable

### Author Declarations

This study was conducted with approval from Monash Health Human Research Ethics Committee (Reference: RES-19-0000333L-53611) and the University of Melbourne Office of Research Ethics and Integrity (Reference: 2024-28658-48562-1). Individual consent was not required as only anonymized data was used.

